# Evaluating the impact of training tools on health workers during the introduction of Pneumococcal Conjugate Vaccine: A mixed method study

**DOI:** 10.1101/2025.08.15.25333756

**Authors:** Abida Sultana, Imkongtemsu Longchar, Guolhoulie Razoukhrielie Rio, Arindam Ray, Amrita Kumari, Seema Singh Koshal, Rhythm Hora, Amanjot Kaur, Rashmi Mehra, Bodhisatwa Ray, Syed F Quadri, Shyam Kumar Singh, Arup Deb Roy

## Abstract

**Introduction:** The Pneumococcal Conjugate Vaccine (PCV) was launched phase wise in India in 2017 and was expanded nationwide in 2021, during the peak of the COVID-19 pandemic. The pandemic restrictions resulted in the hybrid trainings (physical and virtual) for the health workforce. A number of training tools were developed for the health workers to include pre-recorded sessions from immunization experts, presentations and ministry approved Frequently Asked Questions (FAQs), along with the animated videos. As the hybrid model of training was deployed for the first time in the state of Nagaland, the present study aims to evaluate the impact of the training tools used for PCV introduction in Nagaland.

**Methods:** Mixed-method study was conducted in two districts from Nagaland selected using random sampling. In-depth interviews and Focussed Group Discussions (FGDs) were conducted with medical officers, Auxiliary Nurse Midwife (ANMs) and Accredited social Health activist (ASHAs), to gather their experience with the training tools. Quantitative data was collected through a survey among 36 ANMs and ASHAs to evaluate the impact of training tools. Based on Kirkpatrick’s model, qualitative data was thematically analysed using NVIVO 14 software, and quantitative data was analysed using SPSS software v.21.

**Result:** Findings highlight that 29% of the respondents had received training through virtual sessions and 5% via hybrid mode. The majority (66%) of the trainings were in person. Most health workers, particularly the ANMs, found the training materials to be informative and comprehensible. Majority of the respondents preferred in-person training. The finding also revealed the challenges of online trainings included lack of focus, network instability, limited interactions with trainers and clarification of doubts real-time. Furthermore, the videos were highly appreciated for its precise content and comprehensiveness by the respondents (69%). Most of them (87%) found the videos as a pivotal tool for skill-based learning and community sensitization.

**Conclusion:** The study revealed that the field workers still prefer face-to-face trainings. The benefits of online trainings were also highlighted. The training tools had significant positive impacts on health workers’ performance and its effectiveness on community and creating awareness among the communities. The study further makes key program-based recommendations to improve trainings in future. This included sessions with visual aids in the FAQs/leaflets, their availability in local dialects for explicitness and use of more interactive elements such as video-based tools for information dissemination.

## Introduction

Worldwide, pneumococcal pneumonia, caused by *Streptococcus pneumoniae*, is a significant cause of pneumococcal disease-related deaths in children under-five^1^. India is one of the leading countries globally contributing to the pneumonia related mortality rates in under-five children^2^. In 2017, the Ministry of Health and Family Welfare (MoHFW), Government of India (GoI) decided to introduce PCV in Universal Immunization Programme (UIP) after the recommendation of the National Technical Advisory Group on Immunization (NTAGI) in 2015^2^. In March 2020, when COVID-19 pandemic hit the world, the governments across the world took several measures to restrict the spread of this disease^3^. In January 2021, the Government of India launched COVID-19 vaccination campaign to control the pandemic^4^. Simultaneously, in February, 2021, the Indian government announced in the Union Budget that the PCV would be scaled up nationwide using domestic funds considering the burden of pneumonia and the development of the country’s first indigenous vaccine “*Pneumosil*” ^5,6,7^.

As the pandemic restrictions had brought overall health system to a grinding halt impacting majorly on the delivery of public health services, the training of health care workers (HCWs) which is crucial to a new vaccine introduction became challenging as in-person training for PCV introduction was not feasible in many locations^8,9,10^. Therefore, online mode of training model was adopted to capacitate the entire health workers for PCV introduction in the country^11^. In areas with unstable internet connectivity, training was held in a hybrid format (both online and in-person) or conducted entirely in person^11^. For the purpose, a training package was developed which included prerecorded sessions from immunization experts, presentations and ministry approved Frequently Asked Questions (FAQs), along with their translations in regional languages^12^. Furthermore, to mitigate the challenges faced during online training, animated videos, covering different aspects of operationalization of PCV were developed based on the FAQs. These included 5-short videos developed in 13 languages including English, Hindi, specifically for the frontline workers including vaccinators and social mobilizer to reinforce the key information on PCV^11^. The videos were compatible with the smartphones and were circulated to the health workers through WhatsApp.

In the state of Nagaland, PCV was introduced in the month of June 2021^13^. The state has many hard-to-reach areas which are impacted by internet connectivity, and additional challenges (such as security-compromised areas, difficult geographical terrains, etc). In the state too, training of health care workers for PCV introduction followed the hybrid model. This study aims to evaluate the impact of training tools used for training health worker capacity during PCV introduction in Nagaland, a Northeast state of India.

## Methodology

### Study design and Setting

The study utilized a mixed-method approach using both quantitative and qualitative methods. Kirkpatrick Model^14^ was (Figure 1) adopted to evaluate the effectiveness of training tools for Pneumococcal Conjugate Vaccine (PCV) introduction. Descriptive study design was used to depict the perceptions of health workers on training tools and its impact.

**Figure 1:**
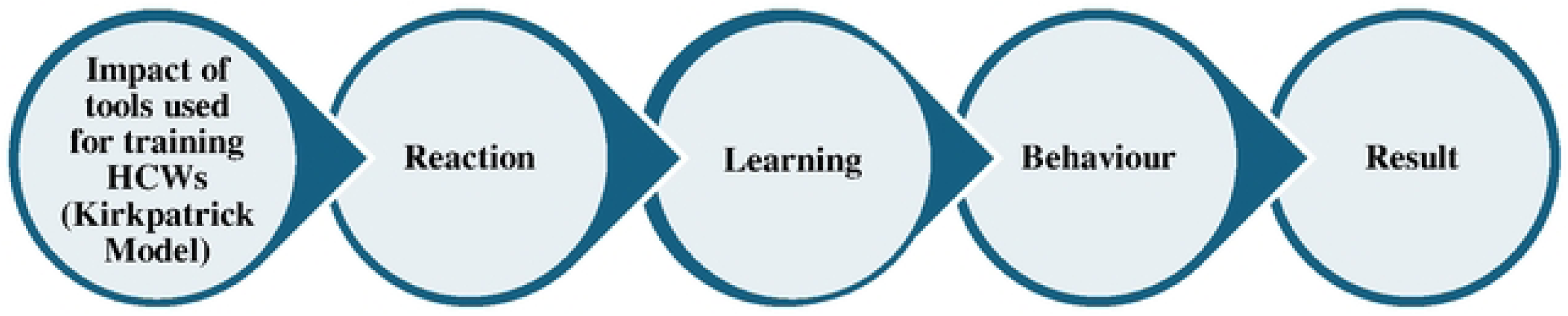
Conceptual Framework of Kirpatrick Model

**Figure 2:**
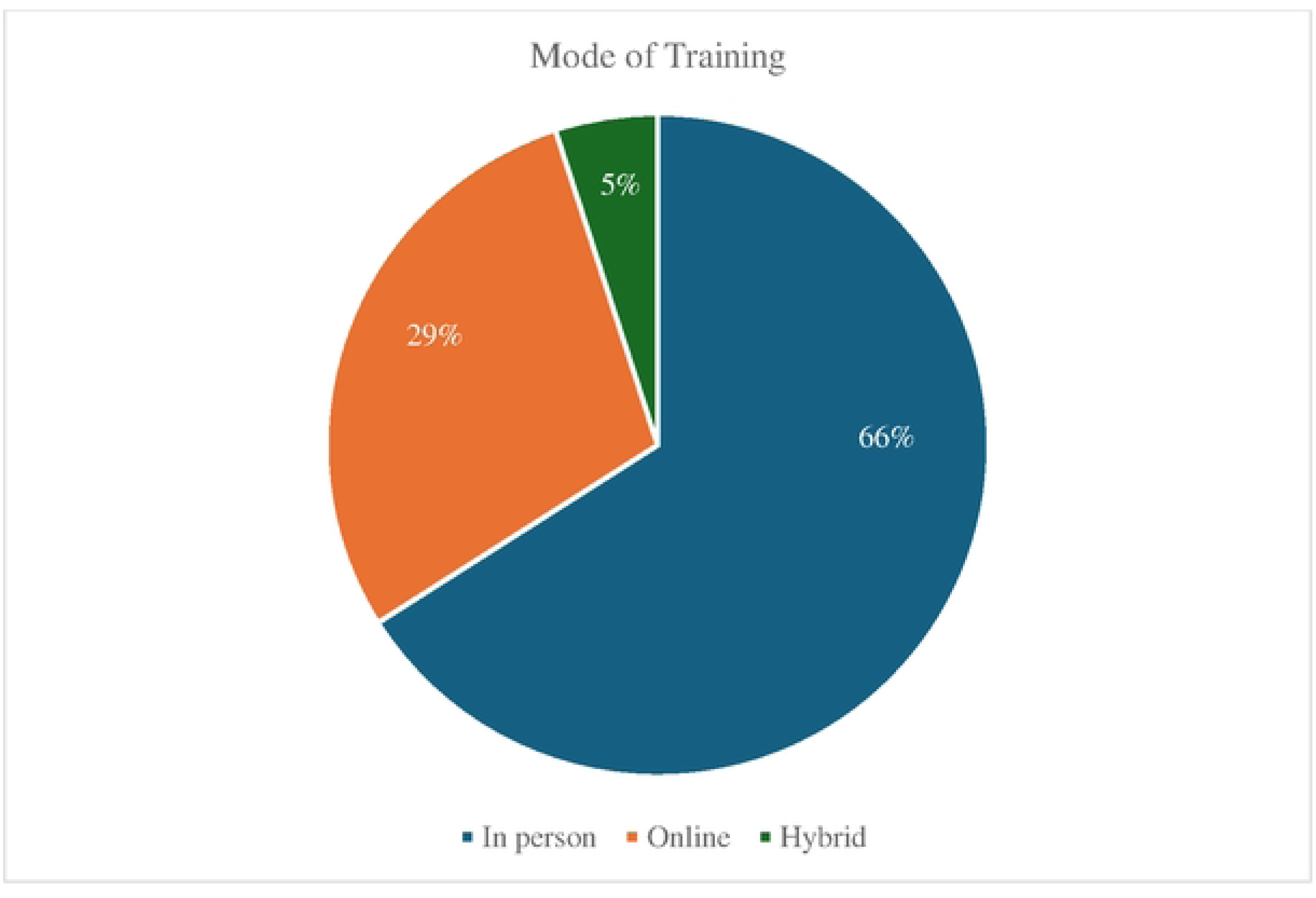
Percentage of HCWs who received training through different modes

The study was conducted in Nagaland, India, from December 14th to 16th, 2022. The state was chosen due to its lowest full immunization coverage for children aged 12-23 months, reported at 58% in NFHS-5 (2019-2021)^15^. Nagaland’s unique geographical terrain, along with infrastructural and socio-economic challenges, contribute to these discrepancies. The state’s mountainous regions and frequent natural disasters, such as floods and landslides, create significant barriers to healthcare delivery^16^. Further, two districts were selected in the state using random sampling technique. Within each district, two blocks were purposively selected in close consultation with the Directorate of Health and Family Welfare, Government of Nagaland (Table 1).

**Table 1:** Geographical coverage for the study.

| State | District | Block |
| --- | --- | --- |
| Nagaland | Dimapur | Nuiland, Urban Dimapur |
|  | Kohima | Kohima Block, Tseminyu |

### Study Respondents and Sampling

For the selection of the health care workers, a list of names and contact details of Auxiliary Nurse Midwives (ANMs) and Accredited Social Health Activists (ASHA) workers who were trained during PCV introduction was taken from block level health personnel of the selected blocks. A sampling frame was generated from the list using simple random sampling technique. The purposively selected respondents hence formed the respondent group for administration of quantitative as well as qualitative tools with respect to ASHA workers and ANMs. Also, for administration of qualitative tool comprising of in-depth interviews, a list of Block Medical officers/ Medical officers (BMOs/MOs) from selected blocks was arranged and target respondents selected based on simple random sampling. Selected respondents were approached by the field teams for taking prior appointments and subsequently questionnaire and discussion guides were administered as per their convenience.

Respondents for the quantitative assessment included ANMs (n=16) and ASHA (n=20) workers while the qualitative assessment included BMOs (n=8), ANMs (n=16) and ASHAs (n=20). The role of BMOs in immunization is to plan, monitor and review immunization sessions within their designated block^17^. The ANMs are responsible for the all the activities related to immunization such as preparing immunization sessions, vaccine administration, and recording, reporting, and tracking of dropouts^18^. The ASHAs are the mobilizers of beneficiaries to the immunization session and they are the primary source of contact with the community^18^.

### Data Collection

For the quantitative data collection, structured survey tools were used while the qualitative data collection was done through in-depth interviews and focus group discussions (FGDs) using semi-structured interview guide. Before the data collection, consent was taken from each respondent for audio-recording the interviews (Table 2). The audio-recordings were later transcribed in verbatim. All the identifiable information of the respondents was anonymized.

**Table 2.** Sample distribution of the respondents included in the study.

| **Approach** | **Respondents** | **Number of respondents per block** | **Total respondents** |
| --- | --- | --- | --- |
| Structured Questionnaire (Quantitative) | Auxiliary Nurse and Midwife (ANM) | 4-5 | 16 |
|  | Accredited Social Health Activist (ASHA) | 5-6 | 20 |
| Focus Group Discussion (Qualitative) | Accredited Social Health Activist (ASHA) | 4-5 | 20 |
| In-Depth Interview (Qualitative) | Auxiliary Nurse and Midwife (ANM) | 4 | 16 |
|  | Block Medical Officer/ Medical Officer | 2 | 8 |

### Data Analysis

The qualitative data was thematically analysed using NVIVO 14 software. Thematic analysis was done using inductive reasoning approach based on the Kirkpatrick model themes of Reaction, Learning, Behaviour and Result. This analysis involved thorough reading and re-reading of the transcripts followed by their coding under each theme. On the other hand, for the quantitative part, descriptive analysis of the survey data using SPSS software v.21 under the thematic heads of Kirkpatrick model in the form of charts was used.

### Ethical Considerations

Ethical clearance for conducting this study was obtained from the Institutional Review Board of Sigma, with issue number 10089/IRB/22-23, on December 4, 2022. The study was conducted in accordance with ethical guidelines and was approved by the Health Department, Ministry of Health and Family Welfare, Nagaland. All participants in this study were employees of the Health Department, Ministry of Health and Family Welfare, Nagaland. The purpose of the study was explained to all participants, and signed informed consent was obtained from each respondent prior to data collection. Respondents were also informed that audio recordings would be kept confidential, anonymized, and their identities would not be shared with anyone.

## Results

Table 3 summarises the key characteristics of respondents included for the present study. Findings of the study were explored and grouped under Kirkpatrick model.

**Table 3.** Demographic characteristics of different respondent categories.

| Demographic characteristic |  | Respondent Category |  |  |
| --- | --- | --- | --- | --- |
| Categories |  | ANM (n=16) % | ASHA (n= 20) % | BMO (n=8) % |
| Age | 20-30 years | 10 | 4 | 2.5 |
|  | 31-40 years | 38.75 | 42 | 60 |
|  | 41-50 years | 32.5 | 48 | 35 |
|  | Above 50 years | 18.75 | 6 | 2.5 |
| Experience | Less than 5 years | 15 | 15 | 40.5 |
|  | 5-10 years | 32.5 | 56 | 52.5 |
|  | More than 10 years | 52.5 | 29 | 7 |
| Educational Qualification | Secondary | 11.5 | 31.1 | NA |
|  | Senior secondary | 43.5 | 35.5 | NA |
|  | Graduate | 45 | 33.4 | NA |

### Reaction

The “Reaction” theme captures health workers’ (ANMs and ASHAs) perceptions of the training received for the introduction of PCV.

### Training

All respondents received PCV training, with 66% receiving in-person training, 29% attending online, and 5% engaging through a hybrid mode. The content was considered informative and relevant, enhancing workers’ confidence in promoting PCV and addressing community vaccine hesitancy.

The study revealed a preference for familiar trainers, such as Medical Officers, among health workers when prompted about the choice of trainers. Participants indicated that these trainers facilitated better understanding through interactive sessions.

As one ANM noted, “*This thought never crossed my mind because my incharge gave proper instructions and explanations and also it was much more comfortable. If it was someone else if might have been difficult to ask any questions to them but with our own MO incharge it was easier to talk to*.”

The study also found that most health workers accessed online training sessions via smartphones, primarily due to pandemic-related restrictions. Training materials shared through platforms like WhatsApp groups and training tools such as videos, posters, and booklets were valued for their clarity and appeal, especially for community outreach. Videos were highlighted for their ease of dissemination and effectiveness in awareness building.

The study explored health care workers’ perceptions of the training sessions provided. It was found that the duration of the training sessions ranged from 1 to 4 hours, typically conducted within a single day. Most respondents considered the sessions to be comprehensive, easy to follow, informative, and sufficient in content. However, there was a clear preference for face-to-face training for future sessions, as it allowed for easier interaction, quicker doubt resolution, and practical demonstrations.

An ANM shared, “*It may be different for others, but for me offline is better. Because, if it is in online all the materials are there in the phone but not the hard copy of it, so things might get misplaced. So, if it is offline and any mistake takes place it is easier to undo it by anyone available*.”

The study identified several challenges faced by health care workers during online training. These included network issues, limited smartphone access, and decreased concentration compared to face-to-face sessions. Network problems were particularly problematic for promoting Pneumococcal Conjugate Vaccine (PCV) efforts, as they hindered the downloading of materials and made it difficult to show videos to community members on small mobile screens.

During the study, participants were asked to provide suggestions for improving the training methods and content, the Block Medical Officers (BMOs) emphasized the importance of incorporating local languages in the training tools for the frontline workers such as the ASHAs, who directly communicate with the community.

A BMO highlighted, *“Since, some of the ASHAs are weak in English and Hindi and does not understand English, it would have been better if material was provided by our own dialect i.e. Nagamese.”*

### Training tools

During the study, an effort was taken to gauge health workers’ (HCWs) experience with the training tools developed for PCV introduction. Nearly all respondents confirmed receiving PCV training tools, including videos, FAQs, booklets, PDFs, posters, and banners, in both printed and digital formats. Digital tools were commonly shared via WhatsApp groups, which was appreciated for its quick and accessible format. Most of the training tools were shared with the HCWs by the BMOs after the training while few were provided at the district level.

Regarding the satisfaction of the respondents on the content of the training tools, it was widely reported that the tools were useful for knowledge acquisition, recall, and as reference material in the field. Visual aids such as pictures, illustrations, and videos, were particularly valued for their clarity and effectiveness in simplifying concepts for both HCWs and community members.

The Block Medical Officers (BMOs) highlighted a need to incorporate local languages, such as Nagamese, into the training tools to improve comprehension, particularly for health care workers (HCWs) like ASHAs who faced challenges understanding English.

A BMO noted, “*The materials and video were good, but like I said, if it was local dialogue, it will be more beneficial especially since ASHA are not that much literate and not everyone can understand English.”*

Another BMO added, “*Yeah, it was simple, so it was easy to understand and beneficial. The video was helpful. The only thing is the video was in English and most people don’t understand, so we interpret and make it simpler.”*

The study also found that visual aids, such as FAQ leaflets, presentations, and videos covering critical vaccine-related aspects, were particularly effective in supporting both learning and communication with the community.

An ASHA shared, *“we understand the basics about the videos, when to give, how to give, and how to explain it to the villagers.”*

Health workers, including BMOs, emphasized that videos were particularly engaging, as they provided exhaustive yet straightforward content.

An ANM shared, “*I am getting older so when I see these videos, I understand but keep on forgetting but the information is retained after I see it several times.”*

### Perceptions about different tools utilized for imparting training

Training tools, including Ministry-approved FAQs in both hard (printed) and soft (digital) formats, played a central role in the PCV introduction. Qualitative assessments revealed that both formats were well-received for their comprehensibility and clarity. Survey findings further supported this, with approximately 73% respondents either strongly agreeing or agreeing that the FAQ leaflets were easy to understand (Figure 3).

**Figure 3:**
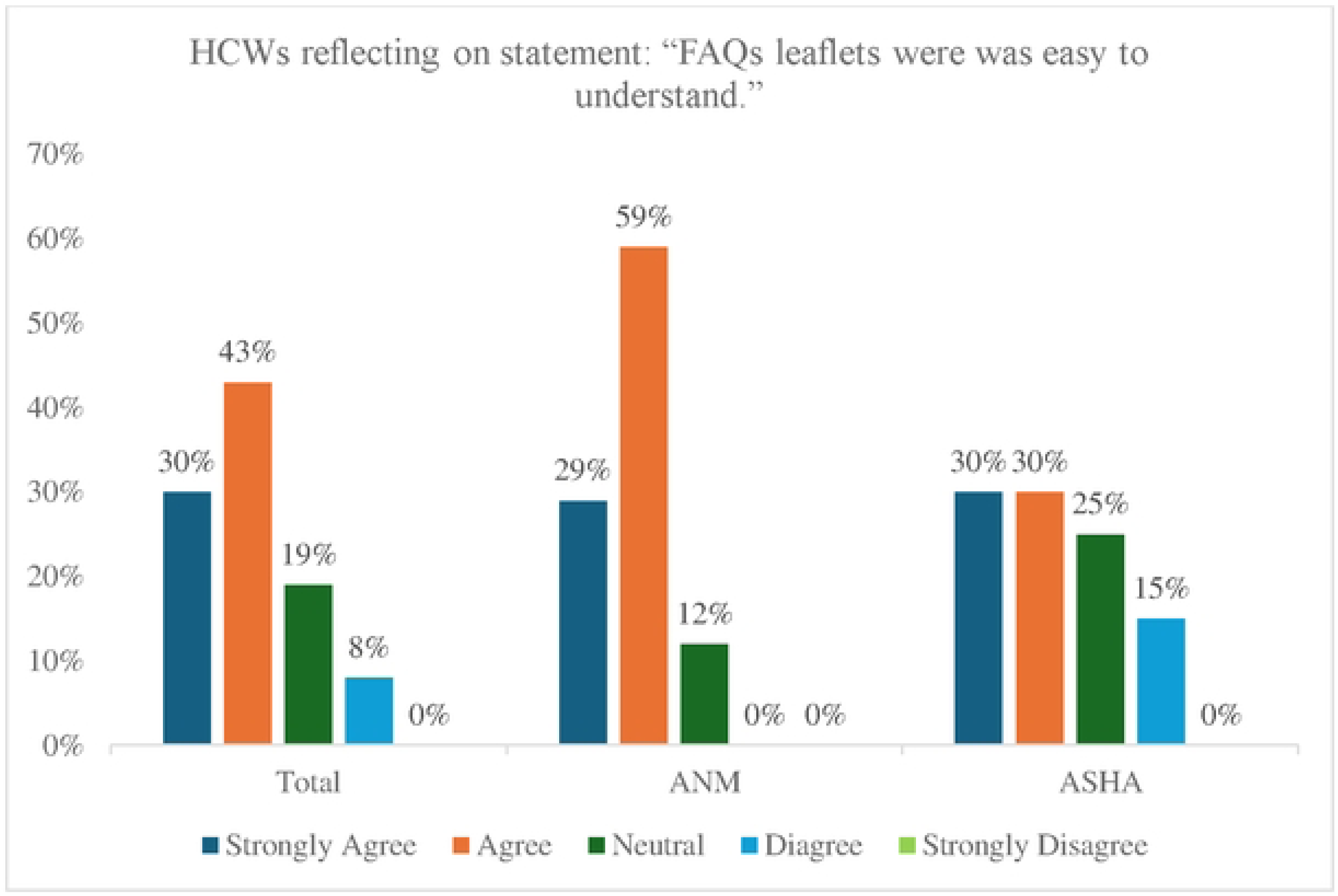
Percentage of HCWs reflecting on statement: “FAQs leaflets/Hardcopy/Soft Copy was simple to understand.”

Similarly, most health workers (HCWs) across various categories found PowerPoint presentations (PPTs) simple to comprehend. Survey results showed that 35% of respondents strongly agreed, and 41% agreed that the PPTs were easy to understand (Figure 4).

**Figure 4:**
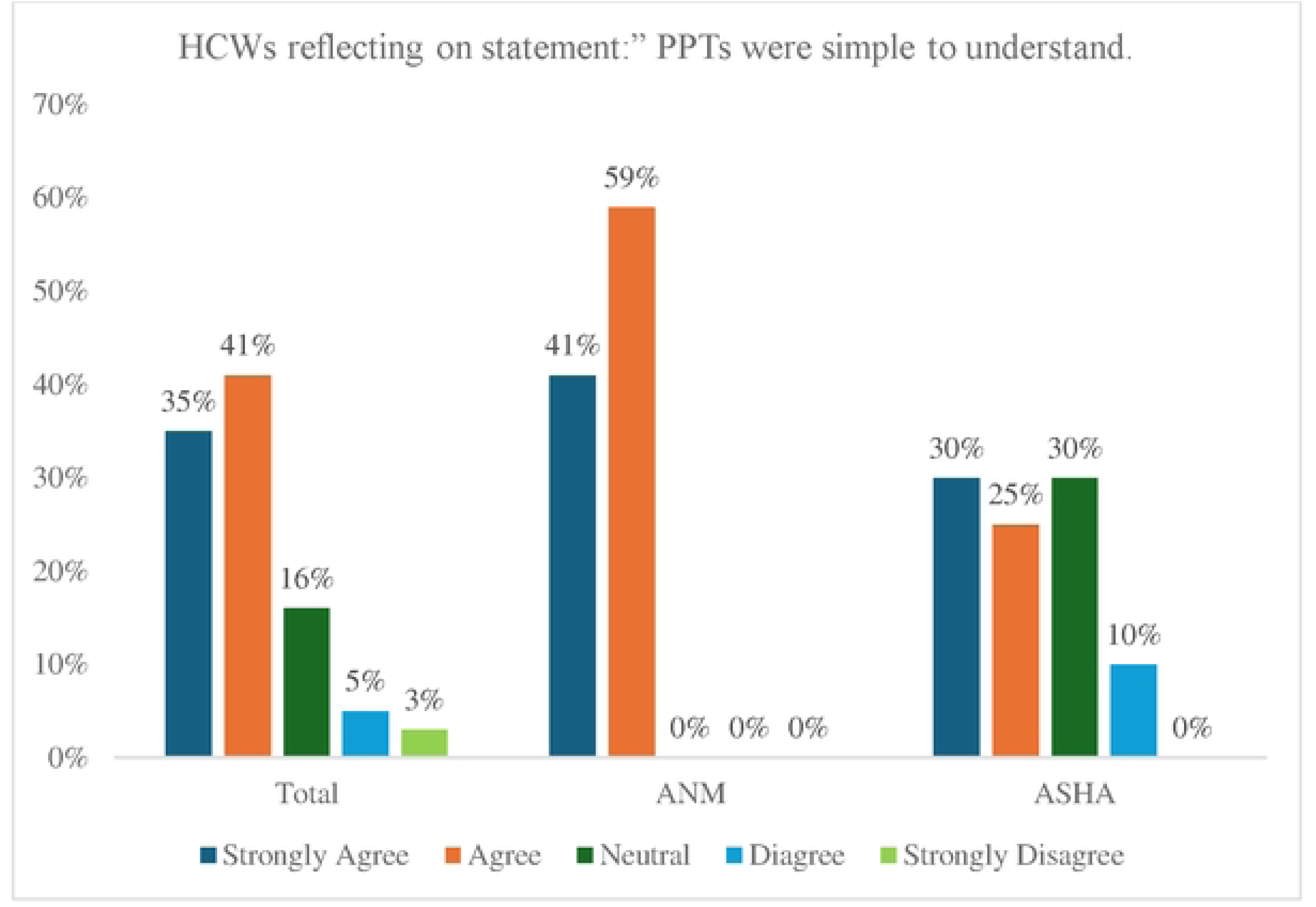
Percentage of HCWs reflecting on statement: “PPTs were simple to understand.”

Frequently asked Questions (FAQ) videos played a significant role in the training tools, enhancing the overall training experience. The Block Medical Officers (BMOs) affirmed that the videos accurately depicted daily scenarios and the populations they serve. Most health workers found the video content adequate for their training needs, with 76% of ANMs and ASHAs agreeing and 63% strongly agreeing. However, approximately 20% of ASHA workers reported difficulties due to the absence of videos in their local dialect (Figure 5).

**Figure 5:**
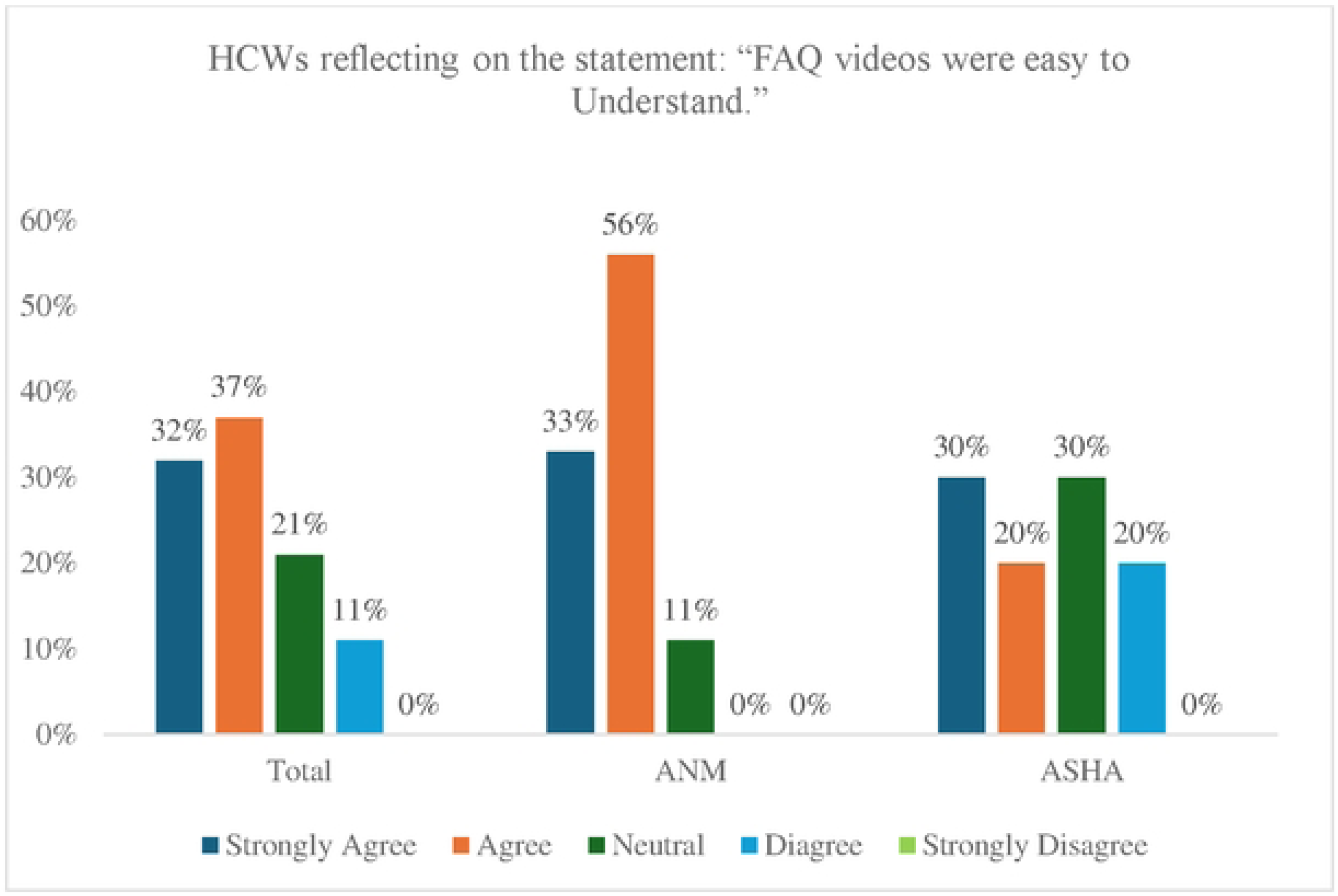
Percentage of HCWs reflecting on the statement: “FAQ videos were easy to Understand.”

Additionally, 36% of respondents strongly agreed and 58% agreed that the videos covered all essential information on PCV introduction (Figure 6). Health workers found the videos to be informative, clear, and skill-enhancing. Moreover, it was noted that HCWs often rewatched the videos whenever clarification was needed.

**Figure 6:**
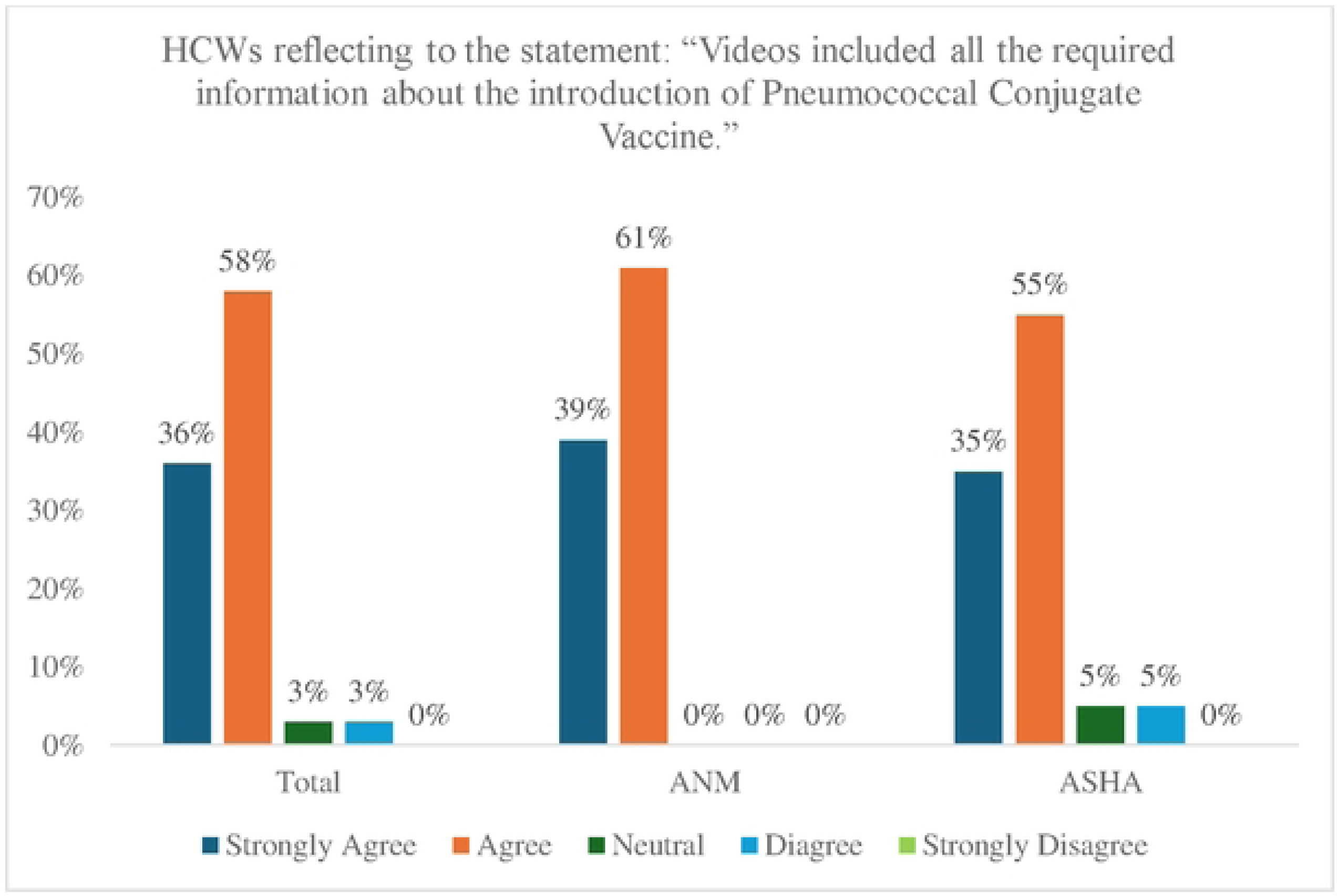
Percentage of HCWs reflecting to the statement: “Videos included all the required information about the introduction of Pneumococcal Conjugate Vaccine.”

While video speed was widely accepted, some viewers noted accent-related comprehension issues. One ASHA shared, *“We are comfortable with English, if we don’t understand the content, we take help of our children.”*

In terms of retention, respondents expressed confidence in recalling video content. Block Medical Officers regularly shared videos with ANMs, mainly through WhatsApp and occasionally email, often resending them for accessibility in case of accidental deletion.

HCWs were also asked to provide suggestions for enhancing the videos in the future. Some suggested that the videos should include more detailed information on the vaccination administration process and be shown on larger screens for better comprehension. Others recommended adding more fieldwork-related content and using local vernacular to improve community connection. Many respondents who related to the animated characters suggested incorporating local dialects for greater impact.

As a BMO mentioned, “*For us, it’s easily understandable but as I’ve said before, it will be good if they also do it in local dialogue, so that everyone could understand, as there are some people who has only passed out class 8 and some are old, so they struggle understanding it all.”*

### Learning

This theme focuses on assessing the learnings of health workers (HCWs) following training sessions, identifying any challenges encountered during the learning process, and evaluating aspects of the training content or process that participants may not have fully understood.

### Contextual and Supportive Training

Among the total respondents, 39% strongly agreed and 61% agreed that the skills learned during training were helpful. However, variations were observed among the HCWs. 44% of ANMs strongly agreed and 56% agreed that the new skills were beneficial. For the ASHAs, 35% strongly agreed, and 65% agreed (Figure 7).

**Figure 7:**
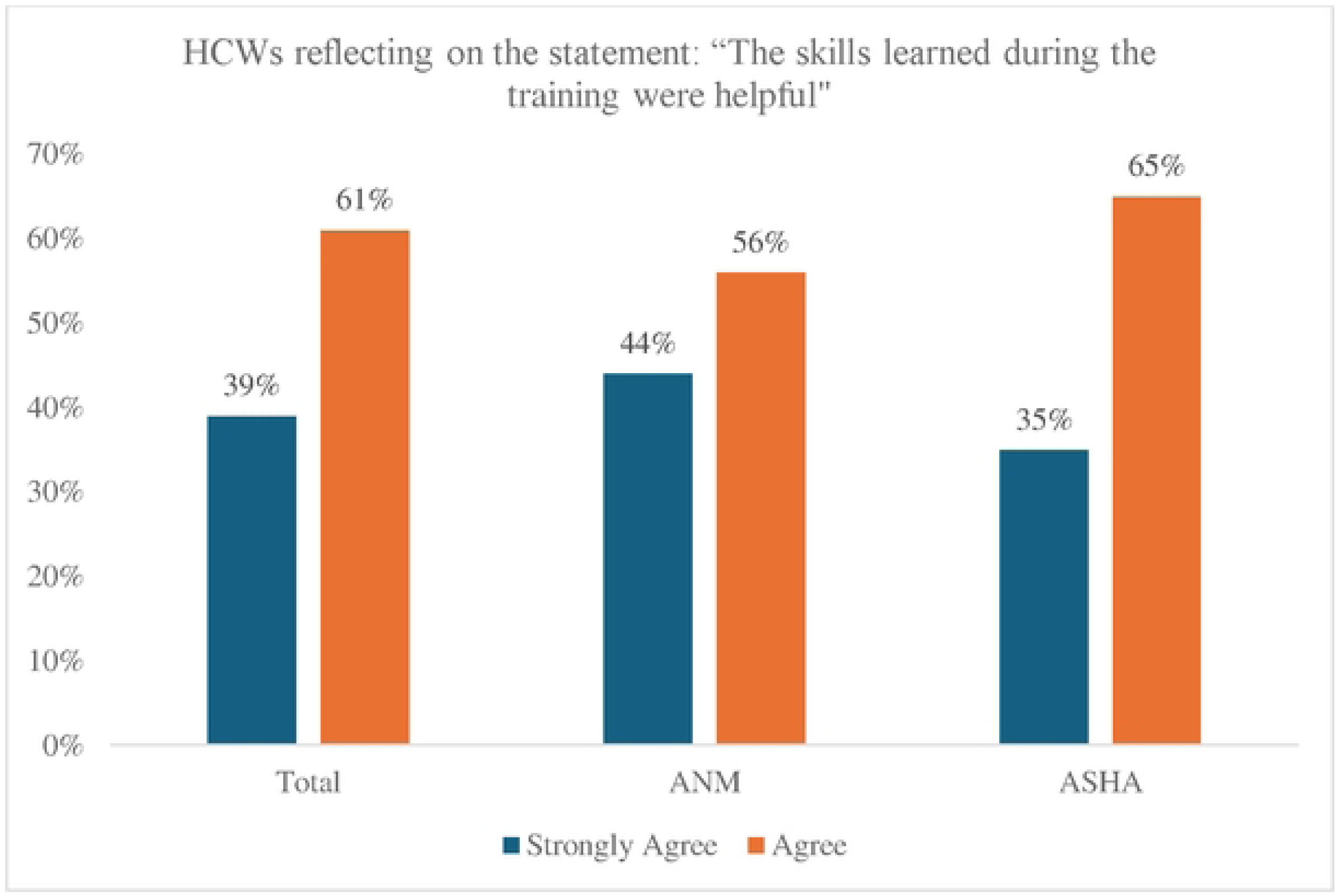
Percentage of HCWs reflecting on the statement: “The skills learned during the training were helpful.”

An ANM stated, “*Earlier, we don’t know anything about PCV before these trainings, hence we came to know about them just last year through trainings and through videos.”*

Videos were recognized as an effective tool for both self-learning and for educating and sensitizing community members. Most of the HCWs agreed that the training videos helped them in disseminating information on PCV to the beneficiaries (Figure 8).

**Figure 8:**
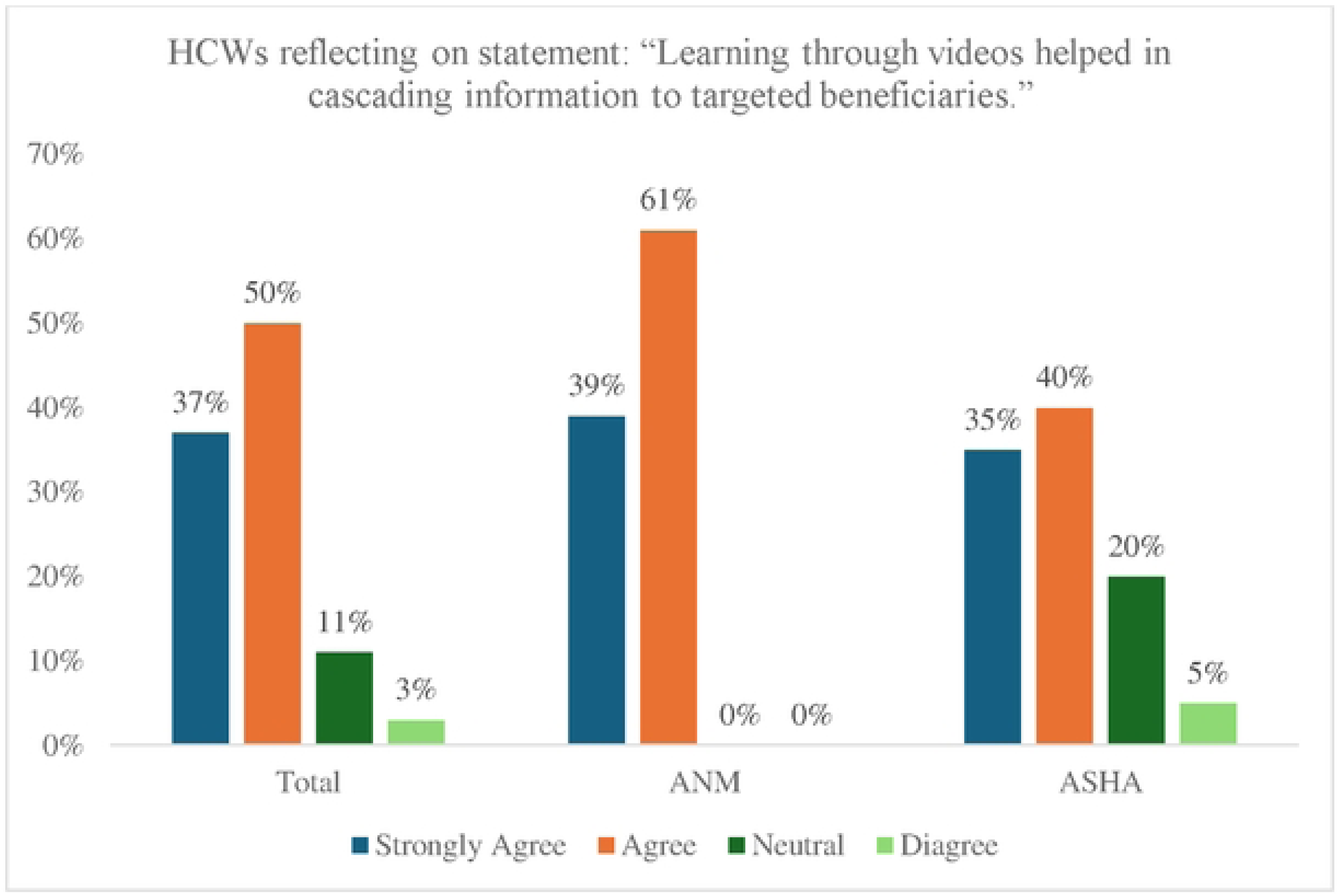
Percentage of HCWs reflecting on statement: “Learning through videos helped in cascading information to targeted beneficiaries.”

When prompted to share key learnings from the training, most of the ANMs highlighted that the training helped them understand PCV and its role in preventing pneumonia in infants. The injection site, dosage, the recommended time interval between doses and awareness of the eligibility criteria for vaccine recipients and information regarding potential side effects were highlighted among some of the key learnings.

### Learning from Training

Most respondents could recall the training content and stated it effectively covered all aspects of the Pneumococcal Conjugate Vaccine (PCV). When prompted about the dosage and site of PCV injection, majority of the respondents were able to accurately answer the questions.

Despite the increased workload, most BMOs expressed confidence in the workforce, stating that ANM and ASHA workers effectively managed the demands. Further, most BMOs felt that the existing training and materials provided were sufficient for frontline workers to enhance service delivery and uptake of PCV.

### Challenges during PCV training and rollout

Most respondents shared positive experiences with the PCV training methods, although some expressed they preferred face-to-face training. They valued the ease of clarifying doubts and improved retention from practical demonstrations. Offline sessions were particularly appreciated for being more interactive, with trainors actively addressing questions and connecting discussions to real-life scenarios.

A few challenges related to the training were noted, including network issues and a need for more detailed content in some of the training videos. Frontline workers (FLWs) found the training videos engaging, clear, and visually appealing. Printed materials were found to be informative ease of reference and could help in vaccine promotion among the community.

### Behaviour

Under this theme, an effort was taken to understand how training has influenced behavior in the context of the application of the learnings to their respective jobs and if there was any change that occurred in their conduct after the training.

### Application of learnings from training into work practices

Most respondents felt well-informed and confident after PCV training, enabling them to effectively apply their learnings in their work. Although, initially nervous, health workers, particularly ANMs reported gaining confidence as they began administrating vaccinations. They noted that the trainings helped create a fully informed workforce of ASHAs. Additionally, ASHA workers utilizing training materials, such as pamphlets and videos, to inform mothers about PCV.

### Application of learning and community acceptance of the vaccine

Most respondents confirmed that training helped them effectively deliver key messages about PCV effectively. Visual aids particularly videos, posters, and pictures, played a crucial role in boosting community awareness and acceptance of the vaccine. Communication materials, including videos and print resources, were found essential in addressing vaccine concerns and persuading parents to vaccinate their children.

### Improvement in skillset

Most ASHA workers reported an improvement in their skills for identifying pneumonia symptoms after the PCV training. They found the training particularly helpful in educating the community and facilitating timely interventions.

Similarly, ANMs indicated that the training videos served as useful guides for the administration of PCV and helped improve their interactions with community members, enhancing their ability to communicate effectively about the vaccine.

## Result

This theme examined the behavioural changes in trained health workers following the PCV training.

It was revealed that most ASHAs and ANMs experienced enhanced work performance post-training, with 34% strongly agreeing and 50% agreeing that their performance improved (Figure 9).

**Figure 9:**
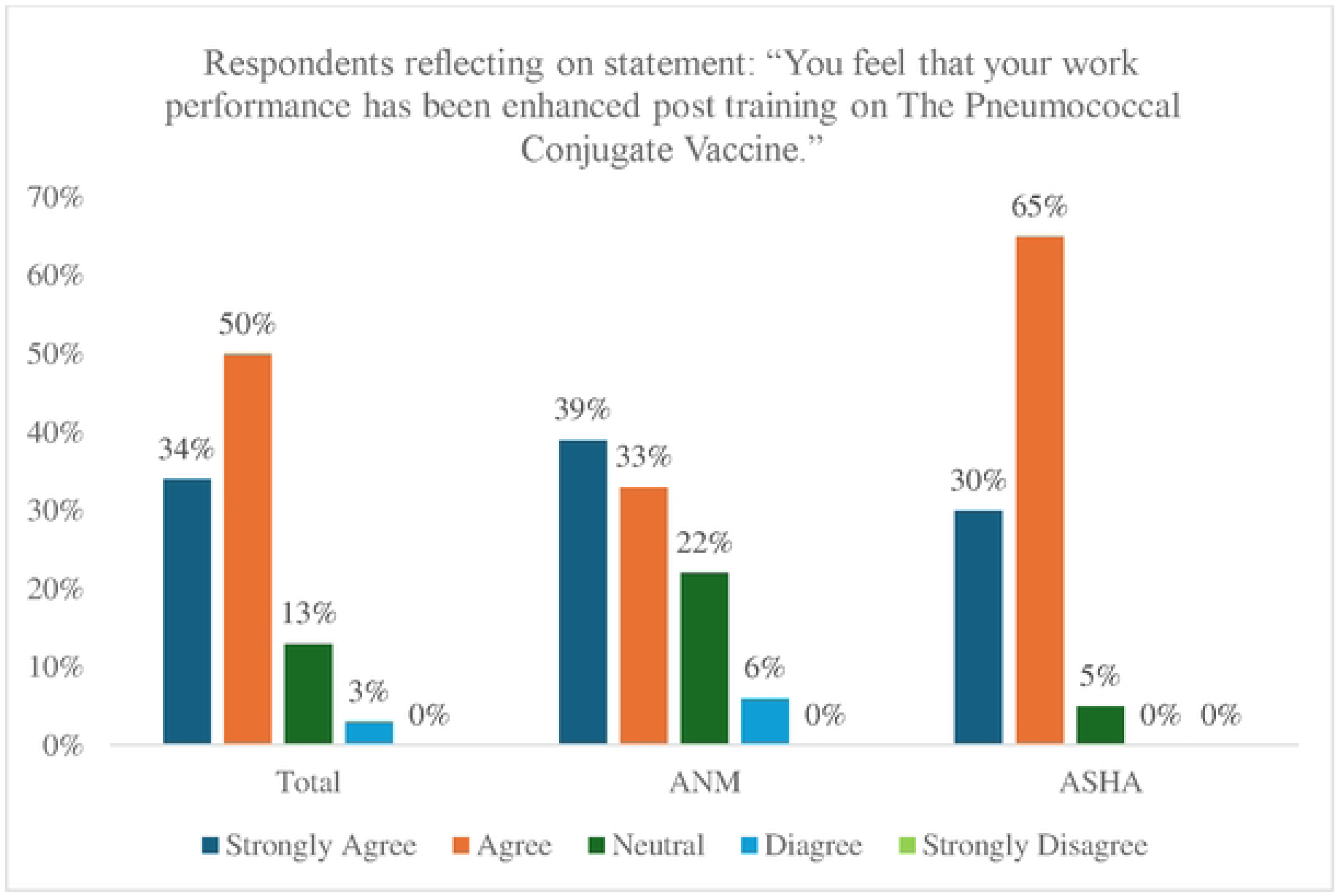
Percentage of respondents reflecting on statement: “You feel that your work performance has been enhanced post training on The Pneumococcal Conjugate Vaccine.”

An ANM mentioned, “*I knew very little about PCV vaccines before I received this training. But after attending this training, my knowledge about PCV vaccines has increased a lot. So, I would say that this PCV training has helped me a lot and made my work a lot easier.”*

Additionally, it was found that most health care workers (HCWs) applied the skills they had learned from the PCV training, with 32% strongly agreeing and 63% agreeing that they effectively incorporated these skills into their work (Figure 10).

**Figure 10:**
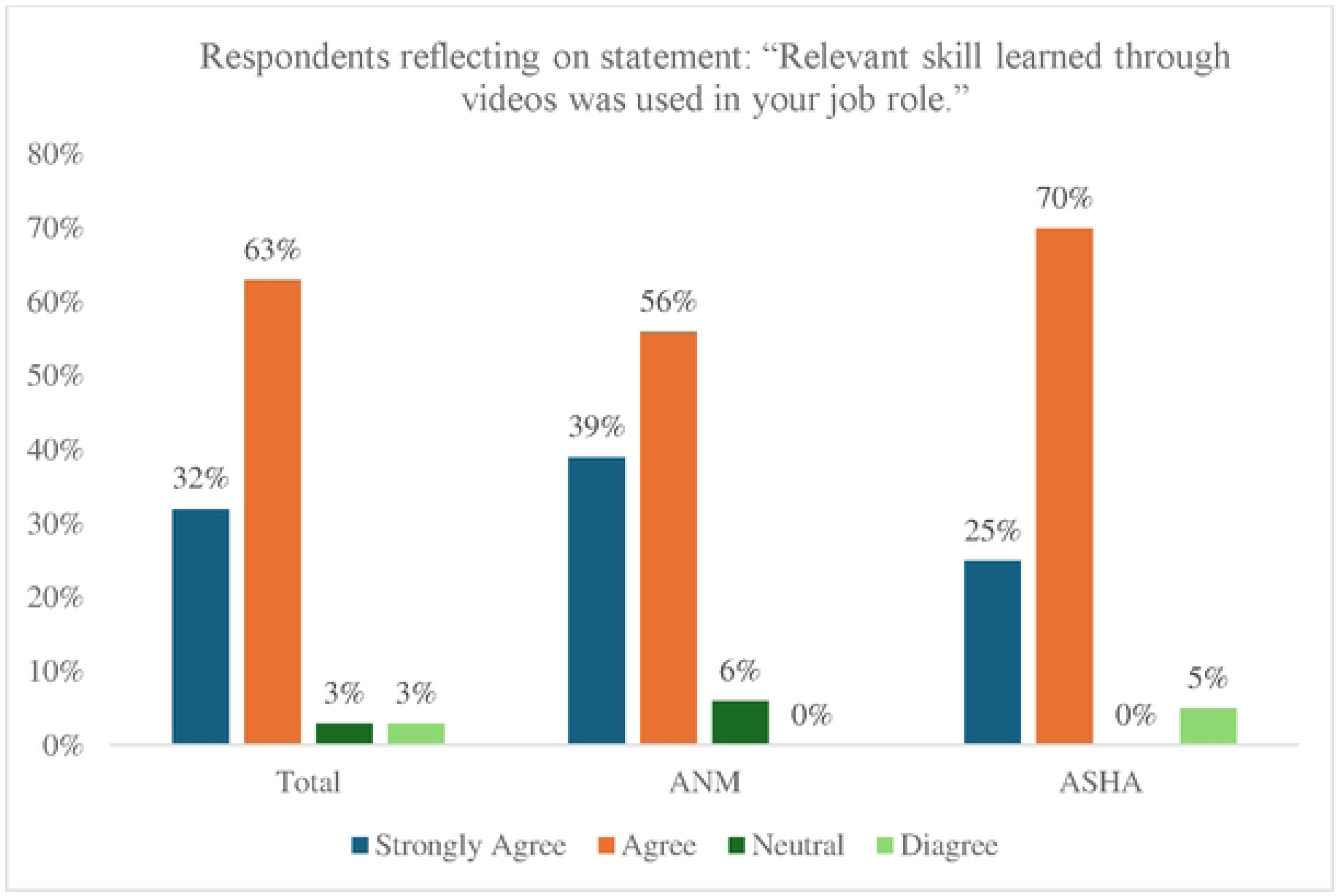
Percentage of respondents reflecting on statement: “Relevant skill learned through videos was used in your job role.”

Respondents also expressed increased confidence in educating the community about the PCV in their respective areas. Approximately 37% strongly agreed, and 61% agreed that their work performance had been enhanced, enabling them to confidently educate and mobilize the community (Figure 11).

**Figure 11:**
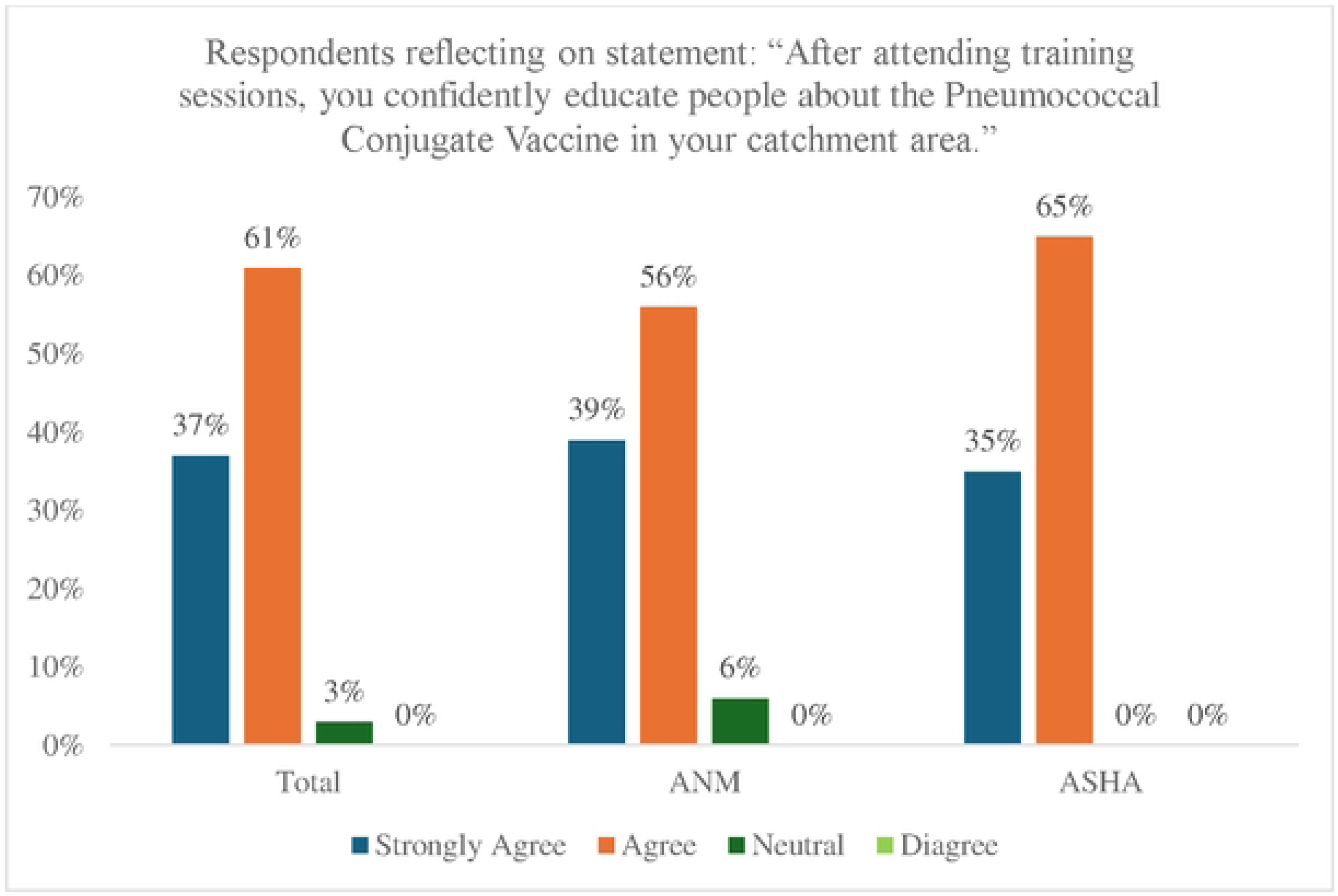
Percentage of respondents reflecting on statement: “After attending training sessions, you confidently educate people about the Pneumococcal Conjugate Vaccine in your catchment area.”

## Discussion

Trainings for the health workers which is a crucial component of any new vaccine introduction were conducted online during PCV introduction in the country due the covid-19 pandemic restrictions. Various training tools such FAQs, booklets, PDFs, videos, posters, and banners, in both printed and digital formats were developed for the health workers and shared with them via smartphones and WhatsApp group. This study was done to evaluate the impact of training tools used for training health worker capacity during PCV introduction in two districts of Nagaland.

The study found that most of the respondents gained confidence after the PCV training allowing them to effectively execute them in their work. The knowledge and the feedbacks from the respondents indicated that the training sessions had been informative and impactful. Similar studies demonstrated positive behavioural changes among respondents after the training, with participants reporting increased confidence in their skills ^19, 20^. Majority of the respondents reported that the training videos helped them gain confidence and enabled them to educate their community about the vaccine in their respective areas. The findings indicated that the training sessions effectively imparted the desired knowledge to the trainees, enabling them to promote the vaccine among beneficiaries and address any vaccine hesitancy. This aligns with the conclusions of Hicks JP et al., who found that Video-based Training and Resources (VTR), facilitated through a digital technology approach, is both a feasible and acceptable method for enhancing clinical knowledge, attitudes, and reported practices^21^.

These training tools were found useful and effective for knowledge acquisition, comprehension, recall and reference. The study found that respondents demonstrated high confidence in recalling video content. The respondents were able to answer any content that they learn from the training tool from their memory and they agreed to the effectiveness of the tools in enhancing their skills and knowledge and disseminating information regarding PCV to the communities. A study conducted by Mahdavi Ardestani SF et al., have also revealed the effectiveness of online learning where the learners can acquire knowledge, learn with flexibility, and have opportunity to review the materials whenever required enabling them to deliver better health services^22^. Another similar finding was consistent in a study conducted by Ajenifuja D eta al, where participants showed positive attitudes and improvements in their knowledge base and changes in their professional practice after online training^23^. Simple and well-organized online learning experiences reduce frustration and maintain learners’ sense of self-efficacy, while focusing on essential content enhances knowledge retention^24^.

The study also found that some health workers were not fully proficient in English and Hindi languages currently used in the materials. It was suggested that incorporating local language into training tools would enhance health workers’ comprehension. This finding aligns with the results of a study by Medhanyie AA et al., where the health workers expressed a preference for mobile health application in local languages, as it would make it easier for them to understand and communicate with the women^25^. Other studies have shown that use of local language have contributed to improved quality of health care service, treatment compliance of patients and improvements of health among patients. When health workers can comprehend, they can explain confidently to the patients leading to better health outcomes^26^.

Despite the benefits of online learning, most health workers expressed a preference for in-person training for future sessions due to ease of interaction, live demonstration, and doubt resolution. Some of the challenges highlighted for virtual training included network instability, less accessibility to smartphones and poor concentration. These drawbacks of online training were consistent with findings from a a study by Bartoletta JJ et al., where respondents reported technical difficulties, poor focus, decrease interaction with virtual learning^27^. Similarly, in a study by Flores MJ et al., respondents from the health sector preferred face-to face training because of limitations of virtual trainings such as poor internet access, scheduling conflicts, and decreased ability to focus ^28, 29^.

However, this finding was in contradictory to another study finding by Becker C et al., in their systematic review of outdoor educational systems, which showed that compared to traditional face to face training models, virtual educational tool had led to improvements in concentration, engagement and retention^30^.

The success of online training during the PCV introduction demonstrates that online training platforms can be leveraged for future vaccine rollouts, particularly in reaching health workers in remote or underserved areas. However, attention must be paid to technological challenges and language preferences to ensure inclusivity and effectiveness. Given the positive feedback and behavioral changes observed, future immunization programs should continue to incorporate training tools that build confidence and empower health workers to educate the public effectively. This is critical for combating vaccine hesitancy and ensuring broad vaccine acceptance. As online and hybrid training models continue to evolve, health authorities can continue to gather feedback from health workers to refine and improve these training tools. Regular evaluation of training effectiveness will help keep strategies relevant and impactful.

There were few limitations to the study. First, the study participants who had received online mode of training represented a small percentage. The responses from the participants are likely to be deflated or inflated as the interview was conducted in English. Another limitation is that the study was conducted post Covid-19 when the trainings have already been completed and therefore is likely to be influenced by recall bias.

## Conclusion

The evaluation of the training tools deployed for Pneumococcal Conjugate Vaccine (PCV) training revealed significant positive impacts on health workers’ performance and confidence. A substantial majority of ASHAs and ANMs reported improved capabilities in educating their communities about the vaccine, demonstrating that the training effectively equipped them with essential knowledge and skills. The comprehensive and interactive nature of the training sessions, along with the availability of engaging informational materials, contributed to enhanced understanding and retention of critical concepts. These findings highlights that the training tools were effective and impactful in fostering desired knowledge and behavioural changes among health workers. Therefore, similar training tools can be utilized for successful introduction and implementation of new vaccines in the future.

## Data Availability

All relevant data are within the manuscript

